# The costs and benefits of an early access scheme for oncology medicines in Ireland

**DOI:** 10.1101/2025.10.30.25339127

**Authors:** Kevin Morris, Mark Pennington, Colette Whitelegg, Luke Parkes, Ronan Mahon

## Abstract

**Background:** Access to new oncology medicines is subject to delays in many countries due to lengthy appraisal processes. This study examines the cost to the Irish healthcare system of implementing a cost-sharing agreement to expedite access to oncology medicines.

**Methods:** The hypothetical cost of implementing an early access scheme in Ireland was estimated for oncology medicines commencing appraisal in 2022. The scheme would have required the manufacturer to cover the cost for the first 180-day period and provide a further rebate if costs per patient over the whole duration of access exceeded those that would have arisen from the finally agreed price. Costs of the new medicine and savings arising from any therapies displaced were estimated on a daily basis using data published in the technical summaries of the National Centre for Pharmacoeconomics, Ireland (NCPE) assessment for each medicine.

**Results:** The scheme would have reduced the time patients waited to access oncology medicines by more than two years on average. Assuming new medicines attract a discount of 30% at reimbursement and that medicines with an existing agreement are subject to a 30% discount, and that a further discount of 10% would be negotiated at reimbursement for the new indication, the costs to the government over the duration of the scheme would have been €61.9m. Earlier access would have generated an additional 1,621 quality-adjusted life years (QALY) over the lifetime of patients accessing the scheme, after discounting. Costs were sensitive to assumptions on discounts negotiated at reimbursement. Costs fell substantially if patients with private insurance were assumed to access care through that insurance.

**Conclusion:** Protracted assessment times lead to substantial health losses to patients with cancer in Ireland. A cost-sharing scheme would accelerate access to new treatments by more than two years and at costs which are unlikely to exceed €62m.

## Introduction

Rapid appraisal of new medicines for inclusion within publicly funded formularies is essential to ensuring timely access to the latest available therapies for patients using publicly funded healthcare systems. Lengthy appraisal processes contribute to suboptimal outcomes for patients, and inequitable access to new therapies if patients with private healthcare are able to access treatment under appraisal by health technology assessment (HTA) bodies. Evidence from Canada indicates that delays in access to oncology medicines can have a substantial impact on patient outcomes with two studies estimating losses of approximately 1,700 life-years associated with delays to access to oncology medicines (1, 2). One of these studies also quantified the impact in quality-adjusted life years (QALYs) reporting a loss of 1,122 QALYs associated with delayed access to nivolumab, afatinib and pemetrexed for non-small-cell lung cancer (1).

Most HTA bodies recognise the negative impact of delays on patients and are committed to providing patients with timely access to new technologies, in principle. However, in practice the HTA process is often delayed. Delays of eight months on average are typical for countries in the European Union (EU) compared with the United States of America (USA) (3). Indeed, research shows that Ireland has one of the longest delays for patients to access new oncology medicines in Europe, with an average of 580 days for all oncology products, and an average of 952 days for combination therapies (4). The Irish Pharmaceutical Healthcare Association (IPHA) has suggested both industry and state need to meet mutual responsibilities to ensure fast and fair access (5). An additional inequity exists in Ireland with the high use of private health insurance (46%) through which patients may be able to gain access to some new oncology medicines (6).

A number of countries have sought to address delays in access to new therapies. For example, the Autorisation d’Accès Précoce (AAP) provides early access to treatments for severe or rare diseases in France (7). Entry to the AAP is conditional on assessment by the Agence Nationale de Sécurité du Médicament et des Produits de Santé (8). In Germany, new treatments are reimbursed following marketing authorisation, with price negotiation commencing after a streamlined assessment period of six months (9). However, a high bar is set for evidence on effectiveness, with randomised controlled trial (RCT) data that includes locally relevant comparators typically being required (9).

Italy operates a ring-fenced budget for innovative medicines that covers the cost of eligible treatments for three years (10).

Furthermore, the Portuguese government has introduced a scheme in which hospital medicines are made available to patients within the public healthcare system as soon as they receive regulatory approval (11, 12). In this scheme, drug acquisition costs are borne by the manufacturer for the first 210 days, and by the government thereafter (11, 12). At the end of the scheme following approval and agreement of a reimbursement price, the manufacturer rebates to the government the costs of drug acquisition in excess of what the government would have paid over the period to that point, if the reimbursement price had been in place at the start of the period (11, 12). In this manner, patients have access to treatment following regulatory approval and the cost to the government per patient is capped at the finally agreed reimbursement price.

This analysis considers the costs of introducing a cost-sharing agreement for oncology medicines appraised in Ireland in 2022, on a similar basis to the Portuguese agreement. The analysis covers the period to the final reimbursement decision for each medicine. The analysis also estimates the additional QALYs accruing to patients through earlier access to oncology treatments.

## Methods

### Cost-sharing structure

The cost-sharing scheme is modelled on the Portuguese scheme (11, 12), amended to a duration of 180 days during which drug costs are paid entirely by the manufacturer. The time period reflects the Health (Pricing and Supply of Medical Goods) Act, 2013, which requires the Health Service Executive, Ireland (HSE) to make a reimbursement decision within 180 days of receiving the application, unless additional data are requested (13). The cost-sharing scheme is assumed to include a preliminary consideration of the clinical data to exclude submissions that would be highly unlikely to be reimbursed. Costs over the first 180 days following the conditional approval date are covered retrospectively by the manufacturer, hence the period of therapy covered by the manufacturer is progressively reduced for those commencing therapy each day after access under the scheme is granted. Costs after 180 days and up until the reimbursement decision are borne by the HSE, but subject to a potential rebate. After conclusion of the HTA appraisal and following agreement of a price, if the total drug acquisition cost paid by the public healthcare system exceeds that which would have accrued with application of the agreed discount, the difference is rebated by the manufacturer. In the situation in which no agreement is reached, the analysis assumes that a retrospective 50% discount to the list price of the drug is applied for calculation of the rebate. The analysis estimates the costs borne by the HSE including the rebate provided by the manufacturer. Figure 1 provides an illustration of cost attribution over the period of the scheme for a single medicine.

**Figure 1:**
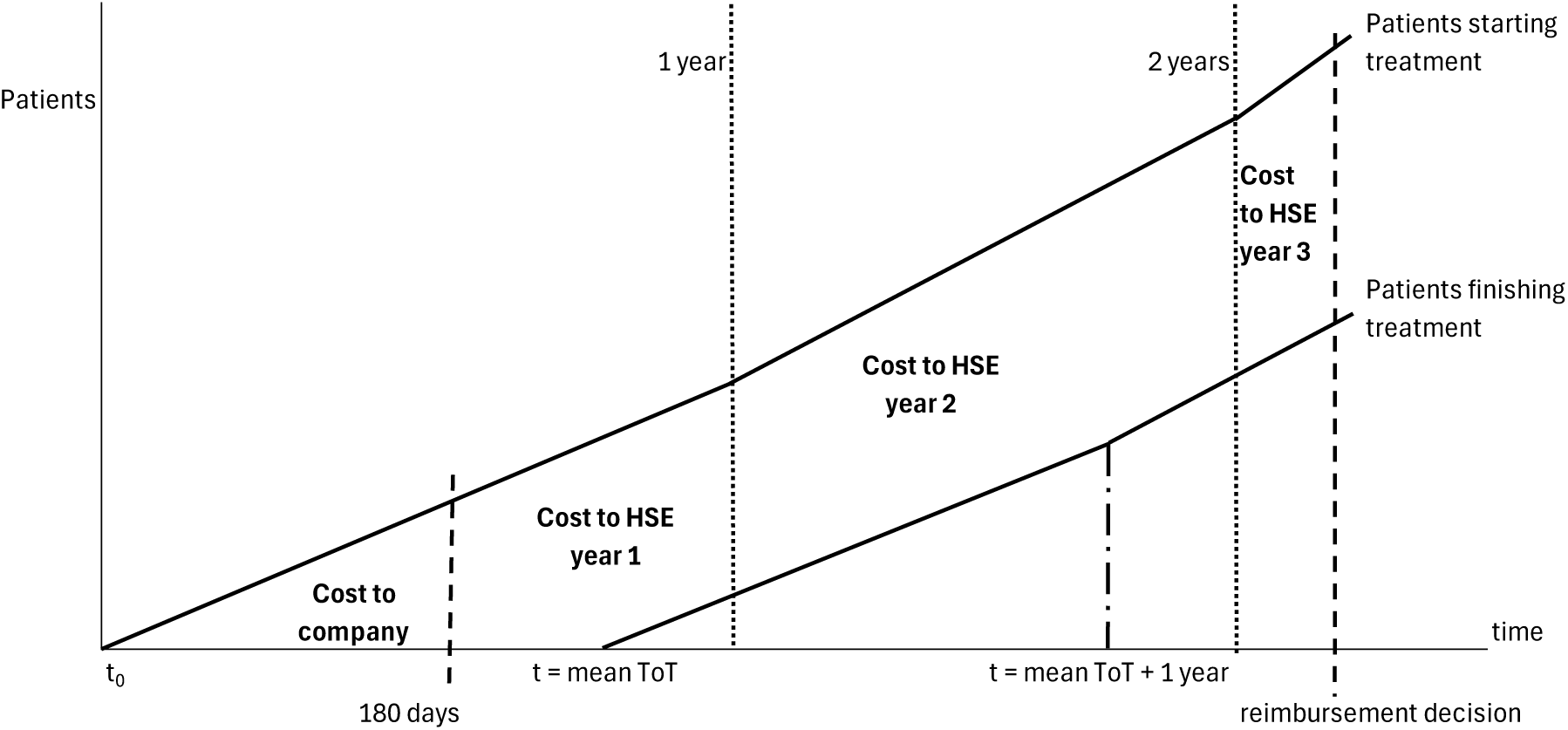
Cost attribution over time for a medicine entering the cost-sharing scheme. Note: diagram represents a medicine with mean ToT < 1 year and with a modest increase in the estimated patient population per year over time. Abbreviations: HSE, Health Services Executive; ToT, time on treatment.

The analysis assumes a regular stream of patients commencing therapy after access is granted and calculates daily costs. The analysis applies the list prices for medicines that are not subject to an existing reimbursement agreement in a different indication. The discounts on drug acquisition costs negotiated by the HSE following approval are not publicly available. The analysis assumes that approval of a new oncology medicine is subject to a discount of 30%. Where a new indication is appraised for a medicine with an existing approval, the pre-existing discount is assumed to be 30% and a further increase to a 40% discount is assumed following approval in the new indication. Our analysis considers only the indication for which each medicine was assessed; in practice any rebate is likely to apply to all licensed indications. Discounts of a further 10% off the list price are considered representative of the likely value of the molecule price discount applied retrospectively across all reimbursed patients over the time of the scheme. These assumptions are tested in sensitivity analysis.

The analysis also estimates the QALYs gained through earlier access to each oncology treatment. Patients starting each oncology treatment are assumed to accrue QALYs equal to the discounted lifetime QALYs associated with the new treatment, minus the discounted lifetime QALYs associated with existing treatment. This difference is the discounted incremental lifetime gain in QALYs associated with the new treatment as reported in the associated NCPE technical summaries. The analysis assumes that patients only commence treatment with the new therapy if it is approved at the point at which they start the relevant line of therapy.

### Sampling frame

The analysis includes all full HTA assessments for oncology indications discussed by the HSE Drugs Group in 2024 (Table 1). Rapid reviews (initial assessment prior to full HTA submission) for these therapies were completed over the period 29/11/2021 to 03/11/2022. The analysis assumes that a conditional approval would be made following the rapid review and that access to the treatment would commence 30 days after completion of the rapid review. The cost-sharing agreement is assumed to be in force until the first of the month in which HSE reimbursement is granted. Table 1 lists the HTA assessments included in the analysis, along with the assumed date of commencement and of termination of the cost-sharing agreement. Two of the oncology submissions appraised by the NCPE were judged to have insufficient clinical evidence to proceed to conditional approval: dostarlimab for mismatch repair deficient/microsatellite instability-high endometrial cancer; and ibrutinib for chronic lymphocytic leukaemia (14, 15). Considerable limitations in the indirect treatment comparisons were noted in the HTA assessments for both dostarlimab and ibrutinib. It is assumed that extension of the rapid review process to consider comparative effectiveness would have identified these limitations.

**Table 1:**
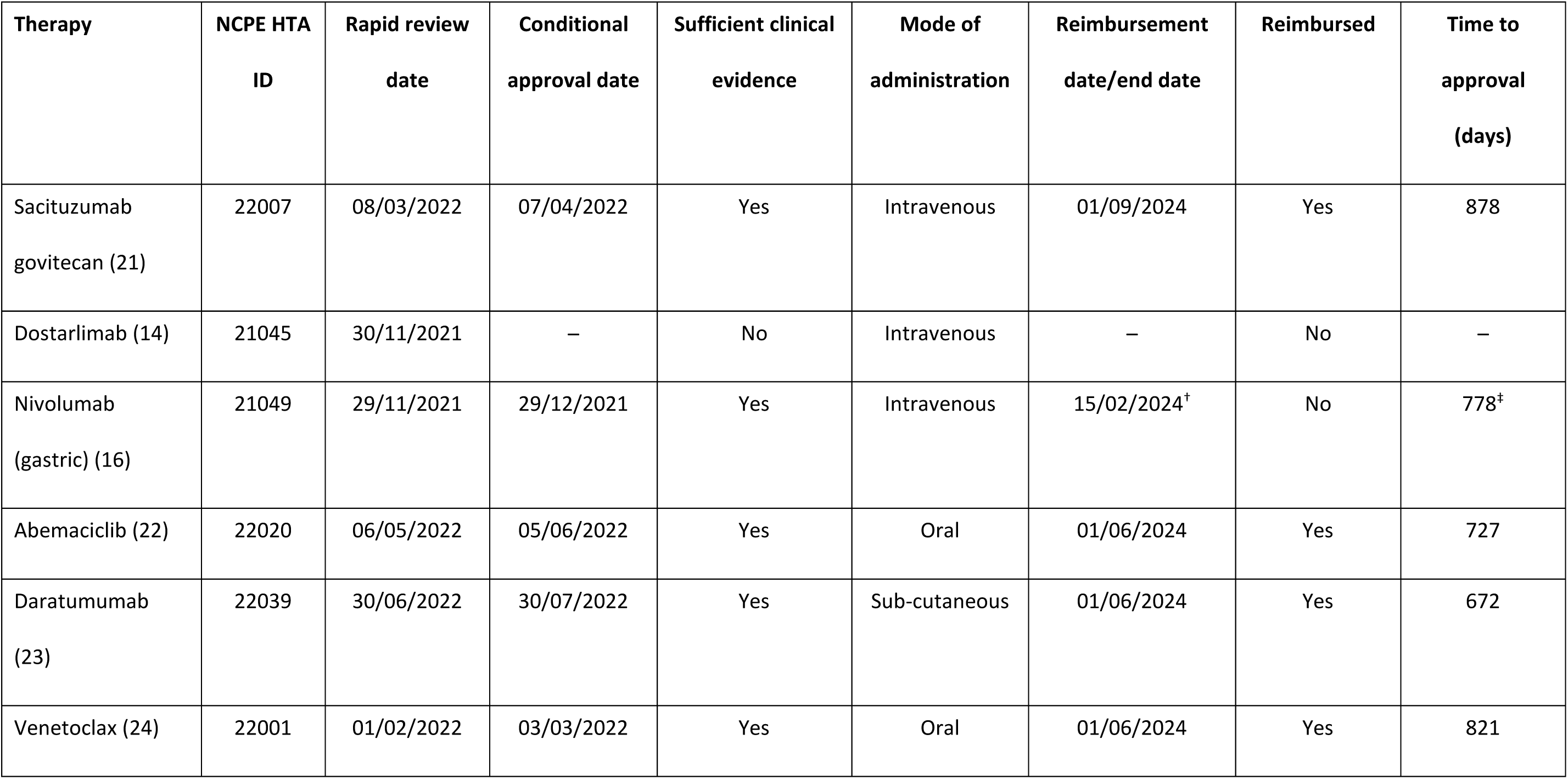
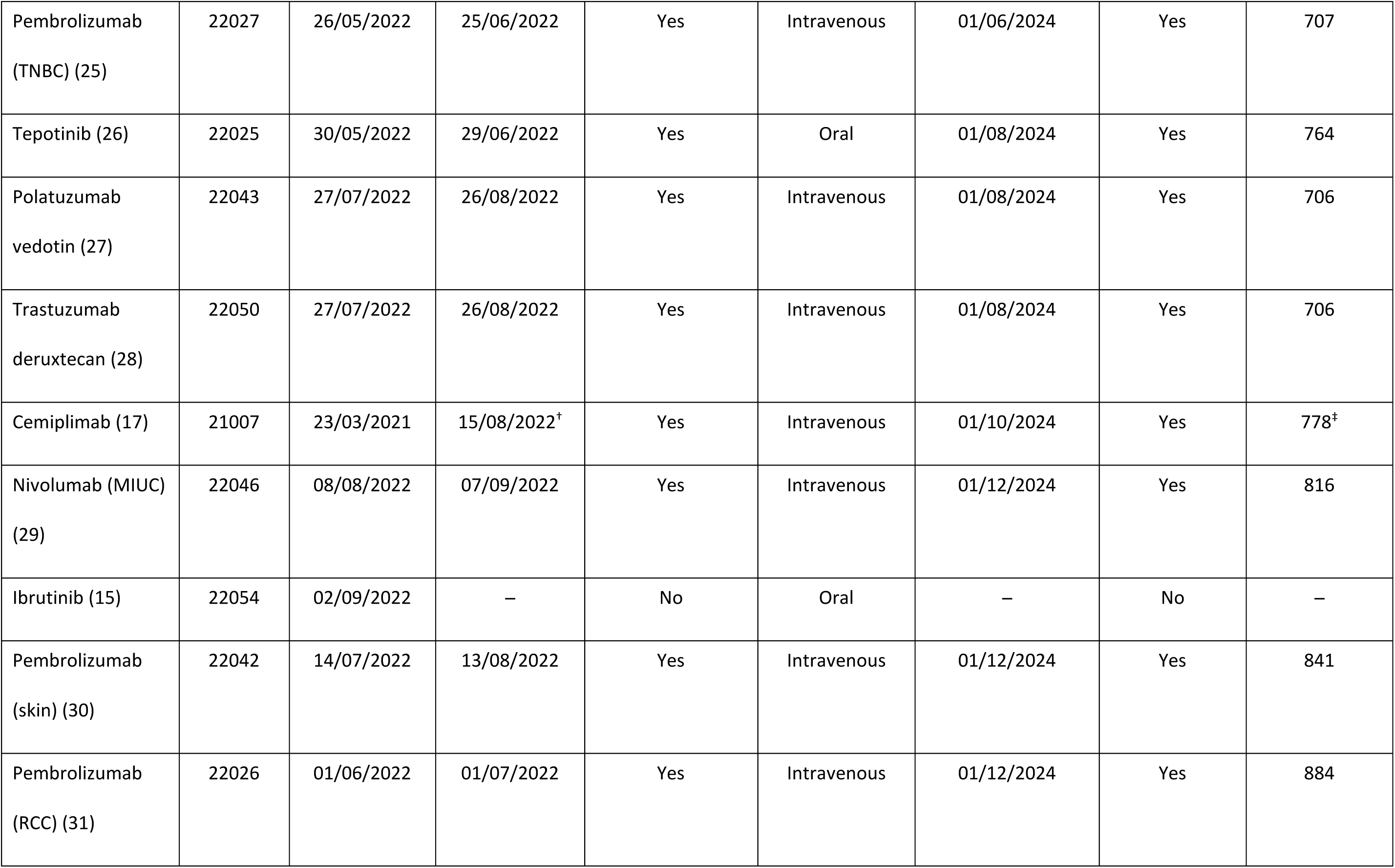
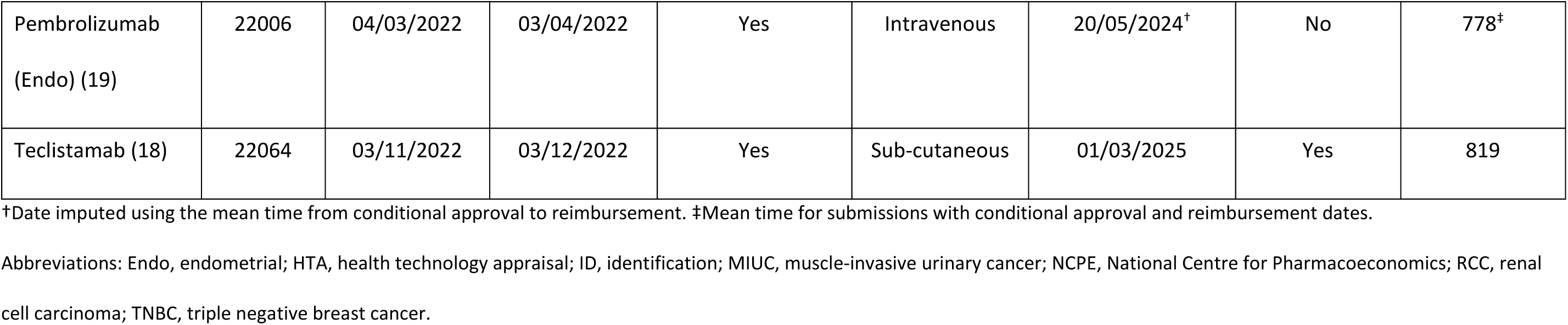
Summary of HTA assessments in Ireland included in the analysis.

The conditional approval dates generated for each of the HTA assessments included in the analysis spanned the period 29/12/2021 to 03/12/2022; there were two days in 2021 in which the cost-sharing scheme would provide access to nivolumab for gastric cancer (16). Costs falling in these two days were included with costs for 2022. There was a considerable delay between the completion of the rapid review and the submission of the full assessment for cemiplimab (17); consequently, a start date for cemiplimab was imputed. The mean number of days from conditional approval to reimbursement was calculated for each of the remaining therapies that received reimbursement. This value was subtracted from the reimbursement date for cemiplimab to estimate a conditional approval date.

Reimbursement dates (and therefore exit from the early access scheme) for all submissions in the analysis fell in 2024, with the exception of teclistamab, which fell in 2025 but are included under costs reported for 2024 for convenience (18). Two submissions were included in the analysis but were not reimbursed following full appraisal: nivolumab for gastric cancer; and pembrolizumab for endometrial cancer (16, 19). End dates for these two submissions were imputed as the conditional approval date plus the mean number of days from conditional approval to reimbursement for reimbursed therapies.

### Data sources

All data for the analysis were sourced from the NCPE technical summaries for each of the oncology submissions. The following data were taken from the technical summaries: gross drug budget impact; net drug budget impact; cost of a course of treatment for the intervention; cost of a course of treatment for the comparator; patients treated in Years 1 and 5, the lifetime discounted QALYs for patients receiving treatment with the intervention and the comparator in the NCPE base case, the total number of patients treated over five years; and time on treatment (ToT). The largest discounted QALYs amongst the comparators were generally selected where there were multiple comparators reported. Two exceptions to this are detailed in the supplementary material.

Where the NCPE estimated higher patient population figures than the company estimates, the analysis applied the midpoint of the two estimates. Where the net drug budget impact was not reported, it was assumed to be the same as the gross drug budget impact. Mean ToT for the intervention was infrequently reported in the technical summaries. Where reported, median ToT was substituted for missing mean. Otherwise, mean ToT was estimated by dividing the cost of a course of treatment by the price to the wholesaler of the intervention therapy, and multiplying it by the duration of a cycle of treatment.

### Analysis approach

Costs were calculated on a daily basis for each therapy from the conditional approval date until the reimbursement date. The analysis assumed that the patient population predicted for each year in the reported budget impact analyses were evenly distributed throughout the year. Duration of treatment with the comparator therapy was assumed to be the same as for the intervention.

Consequently, the number of patients on treatment increased uniformly from the conditional approval date until the mean ToT was reached (assuming mean ToT was less than one year, or no change in patient population in the second year), whereafter the number of patients on treatment remained constant as the number of patients starting therapy equalled the number of patients stopping. Where the technical summary indicated an increase in patient population over time, the number of patients commencing therapy was assumed constant each day within years, with a step change between years. Data on ToT and costs are reported in Table 2.

**Table 2:**
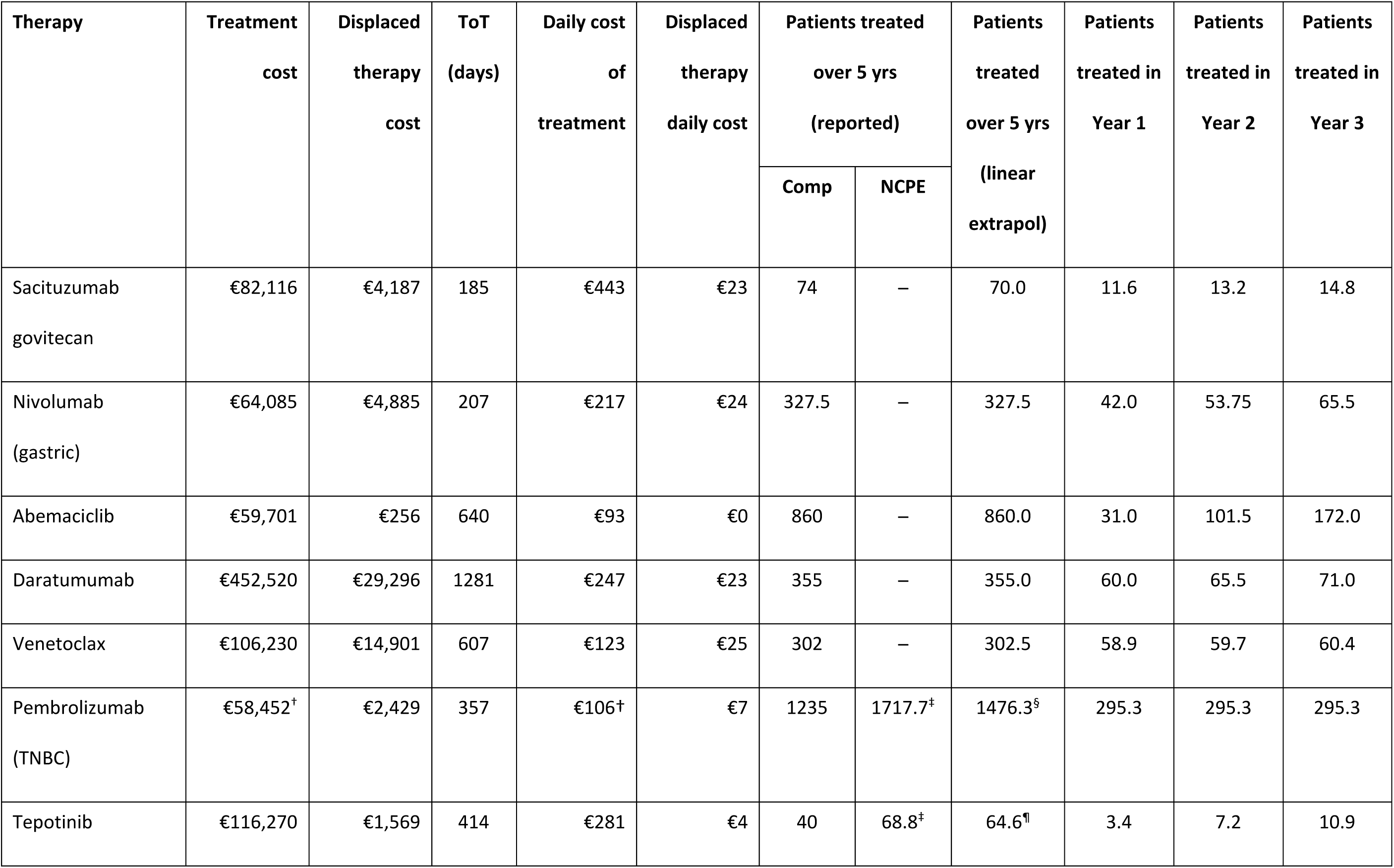
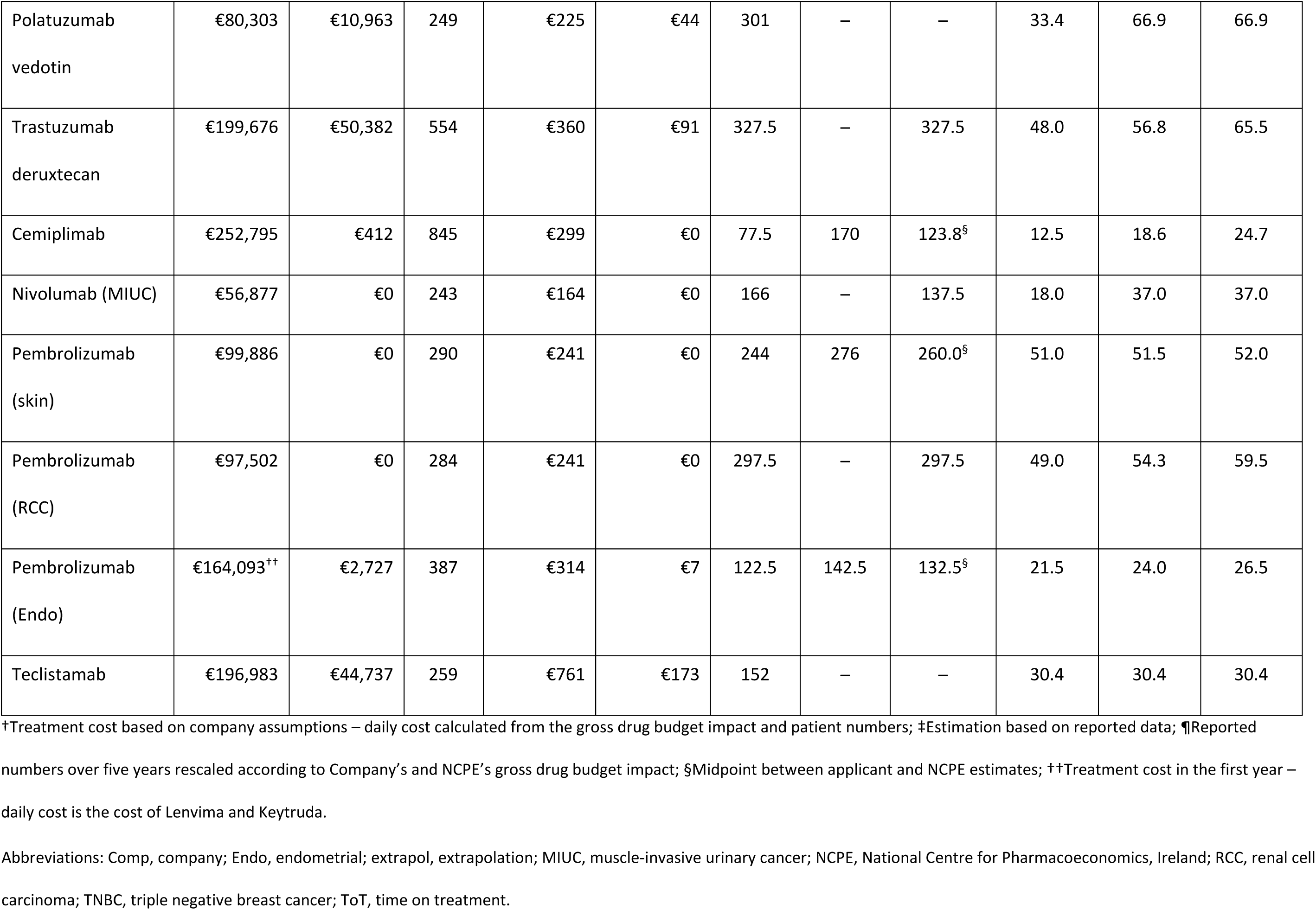
Included therapy costs and patient numbers.

Where patient numbers in Year 1 and Year 5 were reported, a linear increase was assumed to estimate patient numbers in Years 2 to 4. The resulting total patient number using linear interpolation was compared with the reported total patient number over five years. Patient numbers in each year were then scaled such that the calculated and reported patient numbers over five years were the same. Additional details on the calculation of patient numbers are provided in the supplementary material. Patient numbers are reported in Table 2.

A daily cost of drug treatment for each intervention was estimated as the cost of a course of treatment divided by the ToT. This cost was assumed to be accrued on a daily basis for patients receiving the intervention. Likewise, the daily cost of displaced therapies was calculated as the cost of the comparator treatment divided by ToT. Costs per day associated with the intervention and cost savings associated with the comparator could then be calculated by combining daily costs with the total number of patients on treatment in each respective day. Cost savings associated with displaced treatments were assumed to accrue to the HSE throughout the duration of the cost-sharing agreement. Where the cost of a course of treatment with the intervention was not reported the value was estimated as the gross drug budget impact divided by the number of patients treated over five years. Similarly, missing costs for the comparator were estimated as the gross drug budget impact minus the net budget impact, divided by the total number of patients over five years.

Treatment costs were assumed to be constant over the duration of ToT with the exception of costs for pembrolizumab in endometrial cancer (19). Drug treatment in this indication included substantial costs for lenvatinib in addition to pembrolizumab . Consequently, for this submission, a mean ToT was estimated for lenvatinib with pembrolizumab and for pembrolizumab monotherapy, and a daily cost for either the combined treatment or for pembrolizumab monotherapy was applied to patients at the relevant point in their treatment period.

Costs were estimated for 2022 (including 30/12/2021 and 31/12/2021), 2023 and 2024 (including January to March 2025). The following scenario analyses were undertaken: exclusion of patients with private insurance (47.2% held private insurance in 2022 which would have funded oncology treatments delivered in a hospital setting – considered to be those requiring intravenous or subcutaneous administration) (6); exclusion of submissions which were not approved for reimbursement (nivolumab for gastric cancer and pembrolizumab for endometrial cancer); a discount of 20% on the list price for first approvals and 5% for approvals for a new indication; a discount of 40% on the list price for first approvals and 15% for approvals for a new indication; an increase in the period covered by the manufacturer to 210 days; and a reduction in the time taken for appraisals by 180 days.

## Results

The mean time from conditional approval (assumed to be 30 days after completion of the rapid review) and reimbursement was 778.4 days, ranging from 672 to 884 days. Hence, the scheme would have allowed access to oncology therapies over two years earlier than the eventual reimbursement date on average. Table 3 reports the costs falling on manufacturers and the HSE in each of the years 2022, 2023 and 2024, along with the discounted lifetime QALY gains for patients commencing treatment in each of the three years. Costs falling in 2022 are modest and the majority fall on the manufacturer. Costs in 2023 and 2024 are more substantial and fall predominantly on the HSE. Over the three years, costs of €76.9m accrue to the HSE. These costs are offset by a rebate of €15.0m to the HSE, hence the overall additional cost to the HSE is €61.9m. Patients who are able to access treatment through the cost-sharing scheme receive an additional 1,621 discounted QALYs over their lifetime.

**Table 3:** Costs for manufacturers and the NCPE in each of the years 2022, 2023 and 2024.

|  | **2022** | **2023** | **2024** | **Total** |
| --- | --- | --- | --- | --- |
| Company cost | €4,930,259 | €2,077,083 | €0 | €7,007,342 |
| HSE cost | €1,754,606 | €39,873,019 | €35,296,574 | €76,924,199 |
| HSE rebate | – | – | – | €14,990,121 |
| Net HSE cost | – | – | – | €61,934,078 |
| QALY gain | 360 | 785 | 476 | 1,621 |

Abbreviations: HSE, Health Services Executive; NCPE, National Centre for Pharmacoeconomics, Ireland; QALY, quality-
adjusted life year.

Table 4 provides a breakdown of costs falling on the company and the HSE, the number of patients benefitting from earlier access to the therapy, and the number of QALYs gained through earlier access for each of the therapies in the analysis. Typically, costs accruing in the first 180 days were broadly 5–10% of the costs accruing in the subsequent period (to the HSE) before consideration of the costs of displaced therapies and the final rebate to the HSE. Cost savings from displaced therapies varied widely. For three submissions these savings were zero. However, for four submissions (venetoclax, polatuzumab vedotin, trastuzumab deruxtecan, and teclistamab) these costs were more than 20% of the costs of the respective therapy paid by the HSE (after 180 days). Discounted lifetime QALY gains ranged from 10.8 to 605.8 across submissions and were generally reflective of the number of patients accessing the therapy. Box 1 and Box 2 provide detailed examples of the costs and benefits for a medicine that was ultimately reimbursed, and one that was not.

**Table 4:** Costs to company and HSE Table 5: Results of sensitivity analysis.

| **Therapy** | **Cost over 180 days (company pays)** | **Cost from 180 days to reimbursement decision (HSE pays)** | **Costs offset from therapies displaced** | **Rebate to HSE** | **Overall cost to HSE** | **Patients receiving treatment** | **QALYs gained** |
| --- | --- | --- | --- | --- | --- | --- | --- |
| Sacituzumab<br>govitecan | €229,764 | €1,992,196 | €113,295 | €436,824 | €1,442,077 | 30.8 | 10.8 |
| Nivolumab<br>(Gastric) | €406,012 | €3,532,140 | €428,892 | €719,174 | €2,384,074 | 104.3 | 63.6 |
| Abemaciclib | €129,044 | €3,120,590 | €13,924 | €845,846 | €2,260,820 | 131.6 | 89.5 |
| Daratumumab | €661,689 | €8,699,515 | €865,767 | €846,227 | €6,987,521 | 115.0 | 104.7 |
| Venetoclax | €322,073 | €5,921,747 | €1,251,154 | €569,901 | €4,100,692 | 133.5 | 88.1 |
| Pembrolizumab<br>(TNBC) | €1,400,859 | €14,839,558 | €1,038,833 | €919,201 | €12,881,525 | 571.6 | 605.8 |
| Tepotinib | €43,058 | €797,211 | €11,339 | €209,023 | €576,849 | 11.6 | 8.6 |
| Polatuzumab<br>vedotin | €336,197 | €3,769,802 | €800,823 | €416,182 | €2,552,797 | 95.9 | 22.1 |
| Trastuzumab<br>deruxtecan | €771,687 | €11,003,656 | €2,971,121 | €3,141,501 | €4,891,034 | 101.0 | 127.0 |
| Cemiplimab | €166,743 | €3,369,747 | €5,760 | €961,352 | €2,402,635 | 34.4 | 73.2 |
| Nivolumab (MIUC) | €131,269 | €1,911,734 | €0 | €237,174 | €1,674,560 | 63.7 | 86.6 |
| Pembrolizumab<br>(skin) | €548,885 | €6,290,732 | €0 | €639,771 | €5,650,960 | 118.2 | 110.0 |
| Pembrolizumab<br>(RCC) | €526,135 | €6,696,856 | €0 | €505,721 | €6,191,135 | 128.3 | 111.6 |
| Pembrolizumab<br>(Endo) | €301,572 | €4,856,437 | €97,812 | €1,172,145 | €3,586,480 | 49.0 | 46.5 |
| Teclistamab | €1,032,355 | €10,293,131 | €2,572,133 | €3,370,078 | €4,350,920 | 68.2 | 72.9 |
| Total | €7,007,342 | €87,095,052 | €10,170,853 | €14,990,121 | €61,934,078 | 1756.9 | 1620.9 |

Abbreviations: Endo, endometrial; HSE, Health Services Executive; MIUC, muscle-invasive urinary cancer; QALY, quality-adjusted life year; RCC, renal cell carcinoma; TNBC, triple negative breast cancer.

Table 5 summarises the results of the scenario analyses. In the scenario in which the discount negotiated by the HSE is 40% for new therapies and an additional 15% to the existing discount for existing therapies, the rebate increases by a half compared to the base case, and the overall cost to the HSE falls to €47.6m. Correspondingly, assuming a lower discount for first and subsequent indications increases the overall cost to the HSE to €76.3m. Exclusion of the two submissions which were not subsequently reimbursed resulted in a modest fall in the overall cost to the HSE to €56.0m. Reduction in the time to reimbursement by 180 days led to a substantial reduction in costs paid by the HSE prior to reimbursement, and the overall cost to the HSE fell to €39.2m. Extension of the period of time covered by the company to 210 days had a minimal impact on the overall cost to the HSE because reductions in initial costs falling on the HSE were offset by a corresponding reduction in the rebate. Allowing for patients whose care is covered by private insurance generates the lowest overall cost to the HSE of €36.0m.

**Table 5:** Results of sensitivity analysis.

| Scenario | Cost over 180 days<br>(company pays) | Subsequent cost (HSE<br>pays) | Costs offset from<br>therapies displaced | Rebate to HSE | Overall cost<br>to HSE |
| --- | --- | --- | --- | --- | --- |
| Base case | €7,007,342 | €87,095,052 | €10,170,853 | €14,990,121 | €61,934,078 |
| Discount for new submissions/new<br>indications: 40%/15% | €6,345,243 | €79,020,978 | €10,170,853 | €21,276,824 | €47,573,301 |
| Discount for new submissions/new<br>indications: 20%/5% | €7,669,441 | €95,169,126 | €10,170,853 | €8,703,418 | €76,294,855 |
| Exclusion of submissions not approved | €6,299,758 | €78,706,475 | €9,644,149 | €13,098,802 | €55,963,524 |
| Reduction in time to assessment by 180<br>days | €7,007,342 | €53,511,749 | €6,465,939 | €7,826,635 | €39,219,175 |
| Extension of period covered by the<br>company to 210 days | €9,525,510 | €84,576,884 | €10,170,853 | €14,308,947 | €60,097,084 |
| Exclusion of patients with private cover | €3,933,127 | €50,630,454 | €5,972,679 | €8,681,676 | €35,976,099 |

Abbreviations: HSE, Health Services Executive.

## Discussion

This analysis has estimated the costs associated with the implementation of a cost-sharing scheme for oncology therapies subject to full HTA by the NCPE in 2022. Costs extend over a period until March 2025, with costs to the HSE falling predominantly in the years 2024 and 2025. If the scheme were to be adopted, costs would rise over the first three years and then plateau, as medicines exiting the scheme offset the accumulating costs of medicines progressing through approval. Hence the total cost over three years for the therapies considered in 2022 represents a reasonable estimate of costs in each year after the second year. The base case estimate of €61.9m is likely to be an upper estimate of the total costs to the HSE, as it is based on conservative estimates of the discounts negotiated by the HSE. The total costs fall substantially under assumptions of larger discounts. Costs also fall substantially when allowance is made for patients with private health insurance; assuming these patients access hospital administered therapies through their private insurance reduces the cost of the scheme to €36.0m.

The delays to accessing oncology therapies exceeded two years on average for therapies assessed in 2022. This resulted in a loss to patients of 1,621 discounted QALYs over their remaining lifetime.

Hence a cost-sharing scheme would offer a substantial benefit to patients approaching the end of their lives. It is important to note when comparing this figure with the overall cost of the scheme, that costs falling after reimbursement are not included in this analysis. There may be additional treatment costs incurred after the point of reimbursement for patients accessing a new treatment under the scheme. Nevertheless, the patient benefit is substantial when set against the cost of the scheme.

To our knowledge, this is the first attempt to quantify the costs and benefits of an early access scheme for oncology treatments in Ireland. Gotfrit et al, 2020 estimated the impact on patient outcomes of delayed access to treatments for late-stage lung, breast and colorectal cancer in Canada (20). The authors included 21 therapies assessed over the six years from 2011 to 2016, inclusive. The average delay from the first reporting of trial efficacy data to reimbursement was 26.6 months. The resulting loss of access to patients amounted to a loss of 39,067 life years. Assuming each life year has an associated health state utility value of 0.8, the resulting QALYs gained are roughly 20 times the number estimated in this analysis. The population of Canada is roughly seven times that of Ireland and the analysis considers submissions for three common cancers over a six-year period, hence the results are comparable if the three common cancers contribute roughly half of the QALY gains from all oncology submissions. Vanderpuye-Orgle et al, 2022 also estimated life years lost to reimbursement delays for oncology therapies in Canada (1). The authors reported the loss of 1,122 QALYs arising from delays in access to nivolumab, afatinib and pemetrexed for the treatment of non-small-cell lung cancer. Again, considering the difference in the size of the Canadian and Irish populations, this estimate is consistent with our data.

This current analysis has a number of strengths. The sample includes all of the full HTA submissions for oncologic therapies that were discussed in the HSE Drug Group minutes in 2024, and hence are representative of full HTA submissions over a typical year. Data was extracted from the technical summaries of the evaluations of each therapy, hence those data have been subject to detailed external scrutiny in the HTA process prior to publication. Additionally, costs were considered on a daily basis. Finally, two submissions were excluded on the basis that they would have been rejected by a preliminary consideration of the clinical evidence prior to conditional approval, but two other submissions not ultimately reimbursed were included.

The analysis is subject to some limitations. The authors were not privy to discounts in place at the time of rapid review (for therapies submitted for new indications) or subsequently agreed at reimbursement and had to make assumptions. It is evident from scenario analysis that the results are highly sensitive to these assumptions. Some simplifications were applied in the analysis, such as assigning a cost of treatment per day as the cost of a course of treatment divided by mean ToT. Not all the data required were published in each technical summary and sometimes estimations were needed, such as estimating the cost of a course of treatment as the gross drug budget impact divided by the total number of patients treated. No data on the costs of genetic alteration tests that may be required to identify patients for some treatments were reported and hence these costs were not included. The true population for each medicine was assumed as midway between the company and the NCPE estimates, but in many cases NCPE opinion was limited to an observation that the patient population had been underestimated. It is possible that not all HTA submissions in 2022 were discussed by the HSE Drugs Group in 2024 and hence that we did not include all of the relevant submissions.

## Conclusion

A cost-sharing scheme for oncology treatments in Ireland would accelerate access to new treatments by more than two years. The benefits to patients would be sizeable, amounting to 1,621 discounted QALYs for therapies commencing full HTA assessment in 2022. The cost of such a scheme is unlikely to exceed €62m and may be substantially lower.

## List of abbreviations

| Abbreviation | Definition |
| --- | --- |
| AAP | Autorisation d'Accès Précoce |
| EU | European Union |
| HSE | Health Services Executive |
| HTA | Health technology assessment |
| IPHA | Irish Pharmaceutical Healthcare Association |
| NCPE | National Centre for Pharmacoeconomics |
| QALY | Quality-adjusted life years |
| RCT | Randomised controlled trial |
| ToT | Time on treatment |
| USA | United States of America |

## Declarations

### Ethics approval and consent to participate

Secondary data analysis of data in the public domain. Not applicable.

### Consent for publication

Secondary data analysis of data in the public domain. Not applicable.

### Availability of data and materials

The data analysed in this publication are taken from published sources.

### Authors’ contributions

The study was conceptualised by KM. The study was designed by MP, RM and KM. Data collection and analysis was undertaken by LP, CW and MP. MP drafted the manuscript. Editing and manuscript review was undertaken by KM, RM, CW and LP.

All authors read and approved the final manuscript.

## Data Availability

The data analysed in this publication are taken from published sources.

## Acknowledgements

The authors would like to acknowledge Dr Dom Partridge at Source Health Economics for their medical writing and editorial support.

## Supplementary material

Additional information can be found in the supplementary material file provided.

- Selection of the relevant comparator with cost and QALY gains
- Estimation of the population size.

## Box 1: Successful therapy example

### Trastuzumab deruxtecan for unresectable or metastatic HER2-positive breast cancer

The rapid review for trastuzumab deruxtecan for unresectable or metastatic human epidermal growth factor receptor 2 (HER2)-positive breast cancer was completed on 27/07/2022 (HTA ID: 22027). Hence access to trastuzumab deruxtecan would have commenced on 26/08/2022 and covered patients until reimbursement was granted in August 2024 (assumed 01/08/2024). The company estimated that 48 patients would receive treatment in year one, rising to 83 patients in year five. The NCPE did not challenge this estimate. Linear interpolation between the two timepoints provided estimates of 56.8 and 65.5 patients in years two and three, respectively. The cost of a course of therapy including VAT was €199,676 (list price) based on median treatment duration of 18.2 months, which equated to a cost per day of €360. The cost of trastuzumab emtansine displaced by trastuzumab deruxtecan was €91 per day (assumed over 18.2 months). The benefit of treatment with trastuzumab deruxtecan for unresectable or metastatic HER2-positive breast cancer over trastuzumab emtansine was estimated to be 1.26 (discounted) QALYs by the NCPE. Access to the scheme would have provided additional benefits totalling 127 QALYs to 101 patients at a cost of €4.89m to the HSE.

## Box 2: Unsuccessful therapy example

### Pembrolizumab for endometrial cancer

The rapid review for pembrolizumab for endometrial cancer was completed on 04/03/2022, hence access to pembrolizumab (in combination with lenvatinib) would have commenced on 03/04/2022. In the absence of a reimbursement date, a reimbursement decision was assumed on the 20/05/2024, 778 days after access under the scheme. The company estimated that 20 patients would receive treatment in the first year, rising to 29 in the fifth year. The corresponding NCPE estimates were 23 in the first year and 34 in the fifth year. The midpoint of these estimates was taken and linearly interpolated over five years, to estimate the number of patients entering treatment in the first, second and third years (21.5, 24 and 26.5, respectively. Mean ToT for pembrolizumab and lenvatinib was reported as 78.7 and 55.2 weeks, respectively. The cost of a course of therapy with pembrolizumab and lenvatinib over one year was reported as €164,093 (list price) which equates to a cost per day of €314 after applying a 30% reduction. Costs in the second year were reported as €66,633. This cost was assumed to include lenvatinib in the first 21 days of the second year and then pembrolizumab only, generating a cost per day for pembrolizumab only of €243. The cost of chemotherapy displaced by pembrolizumab was €7 per day. The benefit of treatment with pembrolizumab for endometrial cancer over chemotherapy was estimated to be 0.87 (discounted) QALYs by the NCPE. Prior to a decision (not to recommend), 49 patients would have accessed lenvatinib with pembrolizumab, gaining 47 QALYs. The HSE would have paid €4.86m prior to the decision. However, the decision not to recommend would have resulted in a rebate of €1.17m, reducing the overall cost to €3.59m.

