## Supplementary material for "The costs and benefits of an early access scheme for oncology medicines in Ireland"

Supplementary material
   Selection of the relevant comparator with respect to cost and QALY gains

In general, the comparator providing the largest quality-adjusted life year (QALY) gain was assumed to represent standard of care (SoC; as the most effective current therapy. There were two exceptions to this. The comparator for Tepmetko was assumed to be docetaxel rather than nintedanib with docetaxel on the grounds that the technical summary reported a very similar gross and net drug budget impact and the cost of nintedanib is substantially higher than docetaxel. The comparator for Tecvayli was assumed to be treatment of physician’s choice rather than Cilta-cel as the latter drug was under appraisal when the summary was published.

Estimation of the population size

The technical summary did not report the NCPE population estimate for Tepmetko. However, the National Centre for Pharmacoeconomics (NCPE) re-estimated the gross drug budget impact based on increased population estimates. Assuming other parameters were unchanged, the NCPE values for the patient population in each year were estimated by scaling up the respective company values by the ratio of the NCPE’s to the company’s gross drug budget impact.

The number of patients treated in Year 1 and Year 5 was not always reported in the relevant technical summary. Where no indication of the change in population over time was provided, no change was assumed and the population in each year was calculated as a fifth of the total population treated. The respective technical summaries indicated a doubling of patient numbers between years one and two for Polivy for diffuse large B-cell lymphoma and Opdivo for muscle-invasive urinary cancer (MIUC), hence calculations assumed one ninth of the total population treated in the first year and two ninths in subsequent years.

Reporting of the patient population in the appraisal of Keytruda for triple negative breast cancer was particularly opaque. The company estimated that 719 patients would receive neo-adjuvant treatment, and 516 patients would receive adjuvant treatment over five years. The technical summary reported ‘Clinical opinion […] anticipates higher patient numbers (circa 200 patients per year)’. This was interpreted as an NCPE estimate of 1,000 patients receiving neo-adjuvant treatment over five years. The NCPE estimate of the number of patients receiving adjuvant treatment was assumed to be increased by the same ratio as that for the NCPE’s and company’s estimate of patients receiving neo-adjuvant treatment, resulting in an estimate of 718 patients. The true population was then assumed to fall midway between the company’s and the NCPE’s estimate.
